# Exome sequencing reveals aggregates of rare variants in glycosyltransferase and other genes influencing immunoglobulin G and transferrin glycosylation

**DOI:** 10.1101/2022.12.26.22283911

**Authors:** Arianna Landini, Paul R.H.J. Timmers, Azra Frkatović-Hodžić, Irena Trbojević-Akmačić, Frano Vučković, Tea Pribić, Regeneron Genetics Center, Gannie Tzoneva, Alan R. Shuldiner, Ozren Polašek, Caroline Hayward, Gordan Lauc, James F. Wilson, Lucija Klarić

## Abstract

It is often difficult to be certain which genes underlie the effects seen in association studies. However, variants that disrupt the protein, such as predicted loss of function (pLoF) and missense variants, provide a shortcut to identify genes with a clear biological link to the phenotype of interest. Glycosylation is one of the most common post-translationalmodifications of proteins, and an important biomarker of both disease and its progression. Here, we utilised the power of genetic isolates, gene-based aggregation tests and intermediate phenotypes to assess the effect of rare (MAF<5%) pLoF and missense variants from whole exome sequencing on the N-glycome of plasma transferrin (N=1907) and immunoglobulin G (N=4912), and their effect on diseases. We identified significant gene-based associations for transferrin glycosylation at 5 genes (p<8.06×10^−8^) and for IgG glycan traits at 4 genes (p<1.19×10^−7^). Associations in three of these genes (*FUT8, MGAT3* and *RFXAP*) are driven by multiple rare variants simultaneously contributing to protein glycosylation. Association at *ST6GAL1*, with a 300-fold up-drifted variant in the Orkney Islands, was detectable by a single-point exome-wide association analysis. Glycome-associated aggregate associations are located in genes already known to have a biological link to protein glycosylation (*FUT6, FUT8* for transferrin; *FUT8, MGAT3* and *ST6GAL1* for IgG) but also in genes which have not been previously reported (e.g. *RFXAP* for IgG). To assess the potential impact of rare variants associated with glycosylation on other traits, we queried public repositories of gene-based tests, discovering a potential connection between transferrin glycosylation, *MSR1*, galectin-3, insulin-like growth factor 1 and diabetes. However, the exact mechanism behind these connections requires further elucidation.

## Introduction

Genome-wide association studies (GWAS) have so far identified thousands of loci associated with human complex traits and diseases. However, the large majority of these variants are found in noncoding regions of the genome^1^, posing a challenge when attempting to uncover their functional impact on the phenotype. On the contrary, whole-exome sequencing (WES) studies offer the opportunity to identify rare variants of larger effect on the encoded protein, such as predicted loss of function (pLoF) and missense variants, for which causal biological mechanisms are generally easier to elucidate^2^. Methods for exome-wide rare variant analysis have been successfully employed to discover variants and genes associated with both complex molecular traits^3^ and diseases^4,5^. While single-variant tests, such as GWAS, are largely adopted to explore associations of common genetic variants with phenotypes of interest, they have little power to identify rare variant associations, due to the low number of observations. Therefore, a set of methods testing cumulative effects of multiple rare variants in genetic regions, where rare variants are grouped at the gene level (also known as ‘masks’) via a collapsing test, such as burden tests, or variance-component tests (e.g. sequence kernel association test, SKAT^6^) were developed. In addition to increasing the statistical power by aggregating multiple rare-variants, using genetically isolated populations can provide unique opportunities for novel discovery in an association study^7^. Recent bottlenecks, restricted immigration and limited population size lead to increased genetic drift. Consequently, in such populations some otherwise rare variants can substantially increase in frequency compared to the general population, therefore increasing association power for these variants.

Glycosylation is one of the most frequent post-translational modifications, where sugar residues, called glycans, are attached to the surface of proteins. Changes in protein N-glycosylation patterns have been described in the ageing process^8,9^ and in a wide variety of complex diseases, including autoimmune diseases^10^, diabetes^11^, cardiovascular diseases^12^, neurodegenerative diseases^13^ and cancer^14^. Despite glycans having an important role in human health and serving as potential biomarkers in clinical prognosis and diagnosis^15^, we have just started scratching the surface of the complex network of genes regulating protein glycosylation. All studies published to date exploring the genetic regulation of total plasma protein, immunoglobulin G (IgG) and transferrin N-glycosylation have employed single variant-based GWAS tests, mostly uncovering common variants located in non-coding regions of the genome^16–23^. Rare variants contributing to glycan variation, and their impact on human health, thus remain unexplored.

To address this knowledge gap, we used multiple gene-based aggregation tests to investigate how rare (MAF<5%) pLoF and missense variants from whole exome sequencing affect 51 transferrin (N = 1907) and 94 IgG (N = 4912) glycan traits in European-descent cohorts. IgG is both the most abundant antibody and one of the most abundant proteins in human serum. It contains evolutionary conserved N-glycosylation sites in the constant region of each of its heavy chains, occupied by biantennary, largely core-fucosylated and partially truncated glycan structures, that may carry a bisecting N-acetylglucosamine and sialic acid residues^24,25^. Transferrin is a blood plasma glycoprotein that binds iron (Fe) and consequently mediates its transport through blood plasma. Human transferrin has two N-glycosylation sites, with biantennary disialylated digalactosylated glycan structure without fucose being the most abundant glycan attached^26,27^.

In this study, we used gene-based aggregation of rare variants to identify several genes associated with transferrin and IgG glycosylation traits. Significant genes include known protein glycosylation genes as well as novel genes with no previously known role in post-translational modification. Importantly, several associations would not have been detectable by single-point analysis and one association was detected thanks to enrichment of rare variants in population isolates. Finally, we highlight the impact of rare variation in these genes on health-related traits by performing gene-based aggregation tests of 116 health-related traits together with gene lookups in public repositories of gene-based association tests

## Results

### Exome variant annotation

To assess the effect of rare genetic variants on glycosylation of two proteins, we sequenced the exomes of 4,801 participants of European ancestry. After quality control, a total of 233,820 distinct autosomal coding genetic variants were available in the ORCADES cohort (N=2090), 244,649 in the VIKING cohort (N=2106) and 340,203 in the CROATIA-Korcula cohort (N=2872). Percentages of variants for each effect category in the total sequenced coding variation are similar across the three cohorts (Table 1). More than half (∼53%) of the sequenced coding variants are missense variants, of which nearly half (∼28% of total coding variation) are classified as likely or possibly deleterious by multiple variant effect predictor algorithms (see Methods). The second most represented effect category is synonymous mutations (∼33%), followed by variants in splice regions (∼8%), predicted loss of function (pLoF) (∼4%) and inframe insertions/deletions (∼1.5%). Around one quarter of coding variants in the ORCADES and VIKING cohorts are singletons (minor allele count, MAC=1); this percentage is instead higher in CROATIA-Korcula cohort (∼35%), possibly due to the larger sequenced sample size.

**Table 1:** Number of coding exome variants sequenced in the complete sample of 3 isolated cohorts. Counts and prevalence of autosomal variants observed in WES-targeted regions across all individuals in the ORCADES, CROATIA-Korcula and VIKING cohort, by type or functional class for all and for singleton variants (MAC= 1).

|  | ORCADES (N=2090) |  |  | CROATIA-Korcula<br>(N=2872) |  |  | VIKING (N=2106) |  |  |
| --- | --- | --- | --- | --- | --- | --- | --- | --- | --- |
| Variant category | No. of variants | % of total coding variants | Variants % with MAC=1 | No. of variants | % of total coding variants | Variants % with MAC=1 | No. of variants | % of total coding variants | Variants % with MAC=1 |
| coding variants | 233,820 |  | 25.1% | 340,203 |  | 35.5% | 244,649 |  | 28.9% |
| pLOF | 8639 | 3.69% | 37.1% | 12,970 | 3.81% | 47.2% | 9025 | 3.69% | 41.4% |
| Splice acceptor | 872 | 0.37% | 37.8% | 1309 | 0.38% | 45.5% | 945 | 0.39% | 42.7% |
| Splice donor | 1042 | 0.45% | 37.5% | 1506 | 0.44% | 48.1% | 1079 | 0.44% | 40.4% |
| Stop gained | 2833 | 1.21% | 36.8% | 4171 | 1.23% | 47% | 2879 | 1.18% | 41.8% |
| Frameshift | 3401 | 1.45% | 37.8% | 5274 | 1.55% | 47.9% | 3583 | 1.46% | 42.1% |
| Stop lost | 151 | 0.06% | 32.5% | 244 | 0.07% | 43% | 182 | 0.07% | 39% |
| Start lost | 340 | 0.15% | 30% | 466 | 0.14% | 44.2% | 357 | 0.15% | 30.3% |
| Missense | 124,416 | 53.2% | 27% | 183,056 | 53.8% | 37.6% | 130,299 | 53.3% | 30.6% |
| Likely benign (0-1) | 56,366 | 24.1% | 21.8% | 80,777 | 23.7% | 31.6% | 59,141 | 24.2% | 25.4% |
| Possibly deleterious (2-3) | 31,693 | 13.5% | 28.1% | 47,235 | 13.9% | 39.1% | 33,138 | 13.5% | 31.6% |
| Likely deleterious (4-5) | 35,728 | 15.3% | 34.2% | 54,187 | 15.9% | 45.4% | 37,384 | 15.3% | 38.2% |
| Unclassified missense | 629 | 0.27% | 23% | 857 | 0.25% | 29.3% | 636 | 0.26% | 23.1% |
| **Splice region** | 18,580 | 7.95% | 24.7% | 26660 | 7.84% | 32.4% | 19,297 | 7.89% | 27.8% |
| **In-frame indel** | 3383 | 1.45% | 21.5% | 5244 | 1.54% | 29.5% | 3606 | 1.47% | 26% |
| **Protein altering** | 3 | 0% | 33.3% | 4 | 0% | 25% | 2 | 0% | 0% |
| **Stop retained** | 1 | 0% | 0% | 2 | 0% | 50% | 1 | 0% | 25.2% |
| **Synonymous** | 78,798 | 33.7% | 21.1% | 112,267 | 33% | 31.6% | 82,419 | 33.7% | 100% |

### Exome-wide aggregated rare variant analysis of transferrin and IgG glycomes

We performed exome-wide gene-based tests across 51 transferrin traits (glycome subset of CROATIA-Korcula N = 948, VIKING N = 959) and 94 IgG glycan traits (glycome subset of ORCADES N = 1960, CROATIA-Korcula N = 1866, VIKING N = 1086), testing low frequency and rare (MAF <5%) pLoF and missense variants. In total, we identified 16 significant associations for transferrin-(Supplementary Table 1) and 32 significant associations for IgG-(Supplementary Table 2) glycan traits, at Bonferroni-corrected p-values of 8.06×10^−8^ and 1.19×10^−7^, respectively (Figure 1, Table 2). Most gene-aggregated rare variants were associated with protein-specific glycans (transferrin: variants in *FUT6, TIRAP, MSR1* and *FOXI1* genes, IgG: variants in *MGAT3, ST6GAL1* and *RFXAP* genes); only *FUT8* was associated with glycans from both proteins (Table 2, Supplementary Tables 1 and 2). Almost all identified genes encode key enzymes in protein glycosylation (*MGAT3, ST6GAL1, FUT6, FUT8*) or have been previously associated with transferrin and IgG glycan traits in GWAS analysis (*MSR1, FOXI1*)^17,18^. The exceptions are *TIRAP* and *RFXAP*, which have no previously known link to protein glycosylation. We successfully replicated (p-value < 3.2×10^−4^ for transferrin, p-value < 5.9×10^−4^ for IgG) associations of glycans with low-frequency and rare variants from 4 genes - *FUT6* and *TIRAP* with transferrin glycans, and *FUT8* and *MGAT3* for IgG glycans (Table 2) - as frequencies of variants in these genes are similar across the studied cohorts (Supplementary Table 3). While the associations of IgG glycans and variants from *FUT8* replicated, the association of transferrin glycans with variants from the same gene did not reach the significance threshold for replication (p-value in VIKING = 1.7×10^−3^), likely because of the 7-fold decreased frequency of the rs2229678 variant in the VIKING (MAF = 0.0056) compared to CROATIA-Korcula (MAF = 0.049) cohort (Supplementary Table 4). However, given the known biological role of *FUT8* in protein glycosylation as a fucosyltransferase (one of the enzymes involved in the synthesis of glycans), we believe this association to be real. Associations of rare variants from the CROATIA-Korcula cohort in the *MSR1* gene with transferrin glycosylation also did not formally replicate in the VIKING cohort (p-value = 8.6×10^−4^) (Table 2). However, the cumulative allele count of rare variants in this gene is different between CROATIA-Korcula (MAC=46) and the VIKING cohort (MAC=38) (Supplementary Table 3), decreasing the power to replicate. We also detected a couple of isolate-specific associations that are driven by variants increased in frequency compared to publicly accessible biobanks and variant repositories. Namely, the rs750567016 variant in *ST6GAL1* that affects IgG glycosylation is more than 300 times more common in ORCADES (MAF = 3.3×10^−3^) than in UK Biobank (MAF = 1.0×10^−5^) or gnomAD (MAF = 9.0×10^−6^) and is absent from CROATIA-Korcula and VIKING cohorts. The rs115399307 variant in *FOXI1*, associated with transferrin glycosylation, is seven times more common in VIKING (MAF = 2.1×10^−2^) than in CROATIA-Korcula cohort (MAF = 2.7×10^−3^), UK Biobank (MAF = 8.5×10^−3^) and gnomAD (MAF = 7.1×10^−3^) (Supplementary Table 4). While the role of sialyltransferase *ST6GAL1* in IgG glycosylation is well described, the roles of the transcription factor *FOXI1* and the regulatory factor X-associated protein *RFXAP* still need to be confirmed and investigated.

**Table 2:** Gene-based rare variants associations of transferrin and IgG glycosylation.

| Lead glycan | Gene | MAF | Variants | N variants | Discovery cohort | Discovery P | Assoc. test | Discovery MAF | Discovery AC | Repl. cohort | Repl. P | Repl. MAF | Repl. AC | No. of glycans |
| --- | --- | --- | --- | --- | --- | --- | --- | --- | --- | --- | --- | --- | --- | --- |
| **Transferrin** |  |  |  |  |  |  |  |  |  |  |  |  |  |  |
| TfGP20 | *FUT8* | <0.05 | pLoF and deleterious (1/5)* | 6 | CROATIA-Korcula | 6.29x10<sup>-20</sup> | Burden | 0.0111 | 124 | VIKING | 1.73x10<sup>-3</sup> | 0.0042 | 8 | 3 |
| TfGP32 | *FUT6* | <0.05 | pLoF and deleterious (1/5)* | 5 | CROATIA-Korcula | 4.31x10<sup>-18</sup> | SKAT | 0.0097 | 90 | VIKING | 1.56x10<sup>-14</sup> | 0.0072 | 96 | 8 |
| TfGP35 | *MSR1* | <0.05 | pLoF | 3 | CROATIA-Korcula | 6.93x10<sup>-17</sup> | Burden | 0.0083 | 46 | VIKING | 8.64x10<sup>-4</sup> | 0.01 | 38 | 2 |
| TfGP17 | *TIRAP* | <0.05 | pLoF and deleterious (1/5)* | 3 | VIKING | 2.17x10<sup>-10</sup> | SKAT-O | 0.0077 | 44 | CROATIA-Korcula | 8.12x10<sup>-9</sup> | 0.0076 | 98 | 2 |
| TfGP23 | *FOXI1* | <0.05 | pLoF and missense | 3 | VIKING | 1.37x10<sup>-8</sup> | SKAT-O | 0.0074 | 42 | CROATIA-Korcula | 1.56x10<sup>-2</sup> | 0.0014 | 8 | 1 |
| **IgG** |  |  |  |  |  |  |  |  |  |  |  |  |  |  |
| FG2S1/(FG2+FG2S1+FG2S2) | *ST6GAL1* | <0.01 | pLoF and missense | 2 | ORCADES | 9.82x10<sup>-22</sup> | Burden | 0.0019 | 15 | - | - | - | - | 9 |
| Fn/(Bn+FBn) | *MGAT3* | <0.01 | pLoF and deleterious (1/5)* | 4 | ORCADES | 2.31x10<sup>-10</sup> | SMMAT-E | 0.0021 | 33 | CROATIA-Korcula | 6.57x10<sup>-9</sup> | 0.0012 | 29 | 17 |
| FG1n total/G1n | *FUT8* | <0.05 | pLoF and missense | 7 | CROATIA-Korcula | 6.74x10<sup>-8</sup> | Burden | 0.0072 | 177 | ORCADES | 3.04x10<sup>-6</sup> | 0.0037 | 43 | 5 |
| GP21 | *RFXAP* | <0.01 | pLoF and missense | 2 | ORCADES | 1.04x10<sup>-7</sup> | SMMAT-E | 0.0033 | 26 | VIKING | 6.29x10<sup>-2</sup> | 0.0009 | 2 | 1 |
| Lead glycan - glycan trait reporting the strongest rare-variants association at the gene. Gene – gene for which rare variants are grouped. MAF - the highest allele frequency of variants aggregated for the mask. Variants - functional consequence of variants in the mask, aggregated in the given gene (pLoF – predicted loss of function: * denotes the number of algorithms predicting the missense variant to be deleterious). N variants - number of variants included in the mask. Discovery cohort - cohort reporting the lower p-value for the glycan-gene association. Discovery/Repl. P - p-value of the gene-based association with the lead glycan trait in the discovery/replication cohort. Assoc. test – gene-based association test for which p-value is reported. Discovery/repl. MAF - mean minor allele frequency of variants from the mask in the discovery/replication cohort. Discovery/repl. AC - sum of allele counts of variants from the mask in the discovery/replication cohort. Repl. Cohort - cohort reporting the higher p-value for the glycan-gene association. No. of glycans - number of other glycan traits associated with variants from the same mask. Results for transferri glycome are reported at the top of the table, while results for IgG glycome are reported at the bottom of the table. Discovery significance threshold is 8.06x10<sup>-8</sup> for transferrin and 1.19x10<sup>-7</sup> for IgG glycans. Replication significance threshold is 3.23x10<sup>-4</sup> for transferrin and 5.95x10<sup>-4</sup> for IgG glycans. |  |  |  |  |  |  |  |  |  |  |  |  |  |  |

**Figure 1.**
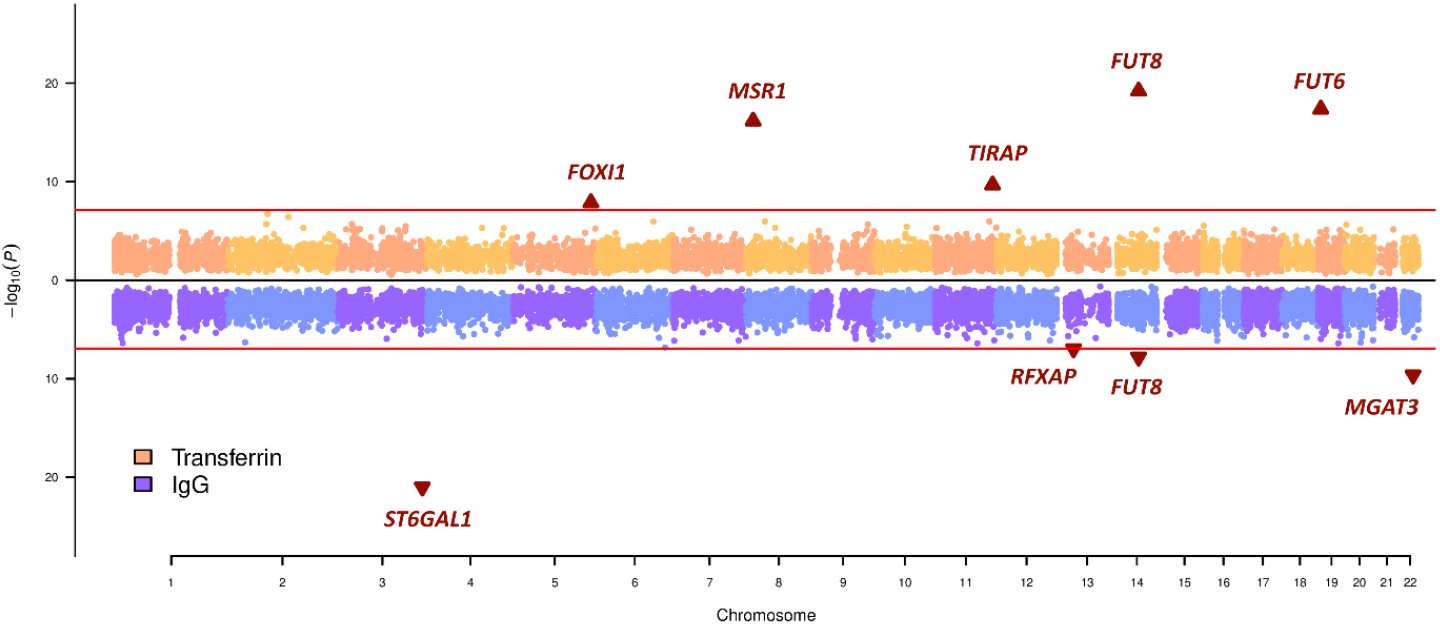
Miami plot summarising the results from exome-wide gene-based tests for transferrin and IgG glycan traits. Genomic positions of the genes, calculated as the mean position of variants included in the reported mask, are labelled on the x-axis and the –log_10_ of the p-value for each rare-variants aggregating test on the y-axis. For each gene-glycan association, the lowest p-value across multiple masks, multiple variant aggregate tests and cohorts was selected for plotting. The Bonferroni-corrected significance threshold for transferrin glycan traits (horizontal red line in the top part of the plot) corresponds to 8.06×10^−8^, while Bonferroni-corrected threshold for the IgG glycan traits (horizontal red line in the bottom part of the plot) corresponds to 1.19×10^−7^. Genes significantly associated with transferrin/IgG glycan traits are indicated with a triangle and labelled, while genes not passing the significance threshold are indicated with dots.

### IgG glycans gene-based aggregation meta-analysis

To further increase statistical power, we performed gene-based aggregation meta-analysis of IgG glycan traits for ORCADES and VIKING cohorts. In addition to two genes already found to be associated with IgG in the cohort-specific analysis (*MGAT3* and *ST6GAL1*), the combined analysis of VIKING and ORCADES cohort added *FUT6* to the list of genes whose rare variants are significantly (p-value<1.19×10^−7^) associated with IgG glycan traits (Supplementary Table 5). *FUT6* is another gene known to be involved in glycosylation^17,20,22,23^, encoding a glycosyltransferase enzyme that catalyses the transfer of fucose moieties to a growing glycan chain.

### Genetic architecture of aggregated effects of rare-variants

To better understand the genetic architecture of identified associations, we next assessed whether our findings could be discoverable within a GWAS framework and whether they are driven by single variants or multiple rare variants working in concert to affect levels of transferrin/IgG glycosylation.

We first performed GWAS on imputed genotypes for each glycan trait and then repeated the rare variant association tests incorporating the dosages of GWAS sentinel SNPs as additional covariates. Genome-wide significant (transferrin p-value<1.61×10^−9^, IgG p-value<2.38×10^−9^) associations reported in this study (Supplementary Table 6 for transferrin glycans and Supplementary Table 7 for IgG glycans) have been described in further details in Landini *et al*.^18^ and Klarić *et al*.^17^. Overall, aggregated associations with variants from 3 out of 8 genes, *FUT8, ST6GAL1* and *MGAT3*, remained significant (p-value <8.06×10^−8^ for transferrin and p-value<1.19×10^−7^ for IgG) after conditioning on sentinel GWAS associations (Table 3). For one gene, *RFXAP*, there were no significant associations in the GWAS analysis, while the remaining four gene-based associations (*FUT6, MSR1, FOXI1* and *TIRAP*) were explained by sentinel GWAS variants. For two of these genes, *FUT6* and *FOXI1*, the GWAS sentinel variants are low frequency (0.02<MAF<0.05; Supplementary Table 8). More specifically, for transferrin glycans, 14 out of the 16 glycome-gene aggregate pairs fail to reach genome-wide significance (p-value <8.06×10^−8^) after conditioning on GWAS sentinel SNPs (Supplementary Table 9), meaning that a considerable part of the rare variant signal was dependent on variants identifiable by GWAS. In contrast, for IgG glycans, 24 of the 32 glycome-gene aggregate pairs remained significant (p-value<1.19×10^−7^), even after adjusting for GWAS sentinel SNPs (Supplementary Table 10).

**Table 3:**
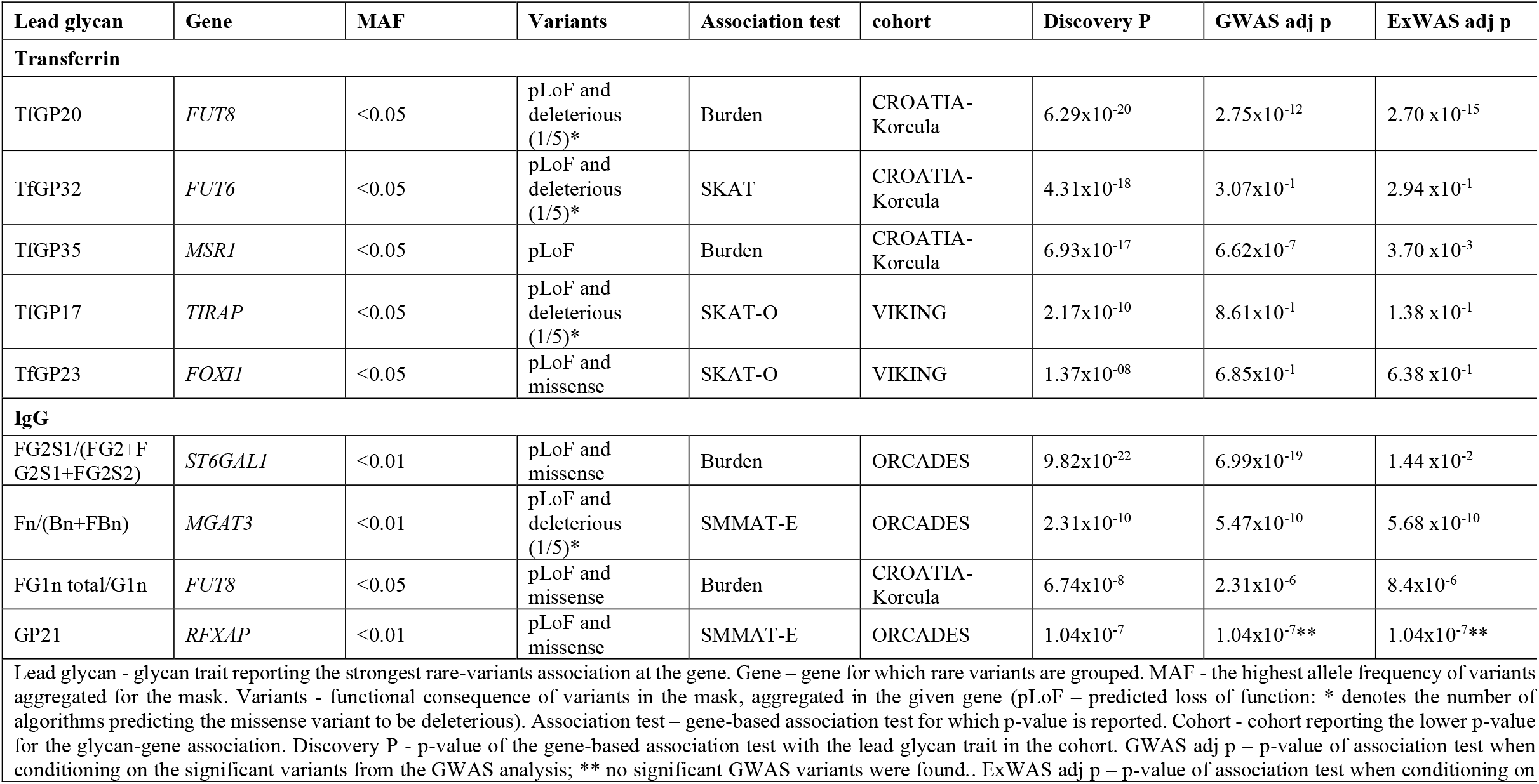
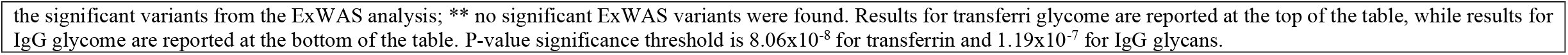
Genetic architecture of aggregated effect of rare variant associations when conditioning on sentinel variants from GWAS or ExWAS analysis. Two associations, those with variants from the *FUT8* and *MGAT3* regions remain significant after conditioning on GWAS/ExWAS sentinel variants. Associations with variants from the *MSR1* gene are dependent on both GWAS and ExWAS sentinel variants. The association with variants from the *ST6GAL1* gene is driven by the sentinel ExWAS variant, which was not present in the imputed GWAS.

| Lead glycan | Gene | MAF | Variants | Association test | cohort | Discovery P | GWAS adj p | ExWAS adj p |
| --- | --- | --- | --- | --- | --- | --- | --- | --- |
| **Transferrin** |  |  |  |  |  |  |  |  |
| TfGP20 | *FUT8* | <0.05 | pLoF and deleterious (1/5)* | Burden | CROATIA-Korcula | 6.29x10<sup>-20</sup> | 2.75x10<sup>-12</sup> | 2.70 x10<sup>-15</sup> |
| TfGP32 | *FUT6* | <0.05 | pLoF and deleterious (1/5)* | SKAT | CROATIA-Korcula | 4.31x10<sup>-18</sup> | 3.07x10<sup>-1</sup> | 2.94 x10<sup>-1</sup> |
| TfGP35 | *MSR1* | <0.05 | pLoF | Burden | CROATIA-Korcula | 6.93x10<sup>-17</sup> | 6.62x10<sup>-7</sup> | 3.70 x10<sup>-3</sup> |
| TfGP17 | *TIRAP* | <0.05 | pLoF and deleterious (1/5)* | SKAT-O | VIKING | 2.17x10<sup>-10</sup> | 8.61x10<sup>-1</sup> | 1.38 x10<sup>-1</sup> |
| TfGP23 | *FOXI1* | <0.05 | pLoF and missense | SKAT-O | VIKING | 1.37x10<sup>-08</sup> | 6.85x10<sup>-1</sup> | 6.38 x10<sup>-1</sup> |
| **IgG** |  |  |  |  |  |  |  |  |
| FG2S1/(FG2+F G2S1+FG2S2) | *ST6GAL1* | <0.01 | pLoF and missense | Burden | ORCADES | 9.82x10<sup>-22</sup> | 6.99x10<sup>-19</sup> | 1.44 x10<sup>-2</sup> |
| Fn/(Bn+FBn) | *MGAT3* | <0.01 | pLoF and deleterious (1/5)* | SMMAT-E | ORCADES | 2.31x10<sup>-10</sup> | 5.47x10<sup>-10</sup> | 5.68 x10<sup>-10</sup> |
| FG1n total/G1n | *FUT8* | <0.05 | pLoF and missense | Burden | CROATIA-Korcula | 6.74x10<sup>-8</sup> | 2.31x10<sup>-6</sup> | 8.4x10<sup>-6</sup> |
| GP21 | *RFXAP* | <0.01 | pLoF and missense | SMMAT-E | ORCADES | 1.04x10<sup>-7</sup> | 1.04x10<sup>-7**</sup> | 1.04x10<sup>-7**</sup> |
| Lead glycan - glycan trait reporting the strongest rare-variants association at the gene. Gene – gene for which rare variants are grouped. MAF - the highest allele frequency of variants aggregated for the mask. Variants - functional consequence of variants in the mask, aggregated in the given gene (pLoF – predicted loss of function: * denotes the number of algorithms predicting the missense variant to be deleterious). Association test – gene-based association test for which p-value is reported. Cohort - cohort reporting the lower p-value for the glycan-gene association. Discovery P - p-value of the gene-based association test with the lead glycan trait in the cohort. GWAS adj p – p-value of association test when conditioning on the significant variants from the GWAS analysis; ** no significant GWAS variants were found.. ExWAS adj p – p-value of association test when conditioning on |  |  |  |  |  |  |  |  |

the significant variants from the ExWAS analysis; \*\* no significant ExWAS variants were found. Results for transferri glycome are reported at the top of the table, while results for IgG glycome are reported at the bottom of the table. P-value significance threshold is $8.06 \times 10^{-8}$ for transferrin and $1.19 \times 10^{-7}$ for IgG glycans.

Next, we performed single-point exome-wide association analysis (ExWAS) and repeated the aggregated rare variant association tests while conditioning on the sentinel ExWAS associations. In this way we tested whether the rare-variants associations with glycosylation were driven by a single variant (i.e. showed an attenuated signal after conditioning on the sentinel ExWAS variant) or were actually affected by multiple rare variants in concert (i.e. associations remain significant after the conditioning). Two of the associations, between variants in the *FUT8* gene and IgG glycans, and variants in the *MGAT3* gene and transferrin glycans, remain significant after conditioning on the sentinel ExWAS variant (Table 3). Upon closer inspection, for both of these genes, the sentinel ExWAS variant was a common variant that is also an eQTL for the gene in blood (eQTLGen^28^: rs35949016, *FUT8* eQTL, p-value = 6.5×10^−159^; rs6001566, *MGAT3* eQTL p-value = 3.9×10^−230^) (Supplementary Table 8). Hence, it appears that glycosylation is affected by common variants and independently by aggregates of rare variants in these two genes. Indeed, by looking at the single-point effects of each rare variant from the mask, we can see that multiple independent rare variants contribute to the effect on glycosylation levels (Supplementary Table 11).

In summary, four of the identified associations, three with low-frequency variants from *FUT6, MSR1* and *FOXI1* and one with a common variant from the *TIRAP* gene, could have been discovered using a GWAS of imputed genotype data. On the other hand, the rare variant association at *ST6GAL1* gene could only have been discovered using an ExWAS, as it is too rare to be imputed well. Finally, associations with variants from two genes, *FUT8* and *MGAT3*, are driven by multiple rare variants simultaneously contributing to glycosylation of IgG and transferrin. Also the rare variant association at *RFXAP* gene could not have been discovered by either GWAS or ExWAS as there were no significant single-point associations. However, it is important to note that we could not replicate this association because the variants from its mask are depleted in the other studied cohorts (Supplementary Table 4).

### Links to health-related traits

We next wanted to assess the potential impact of rare protein glycosylation variants on health. Since some of the gene-glycan associations are population-specific, stemming from the genetic drift in isolated populations, we first performed “gene-level PheWAS” with quantitative health-related traits measured in studied cohorts. At the same time, since these cohorts, because of their sample size, might be underpowered to detect associations with common diseases, we queried public repositories of aggregated rare-variants associations for these genes.

We performed exome-wide “gene-level PheWAS” with 116 quantitative health-related traits measured in the ORCADES, CROATIA-Korcula and VIKING cohorts, limited to the genes containing pLoF and missense variants that were associated with transferrin or IgG glycome variation (Table 2).When possible, we sought to perform the analysis in the same cohort where the glycan-gene association was discovered. The only significant (p-value<5.4×10^−5^) association was with transferrin glycosylation-associated rare variants from the *MSR1* gene and blood levels of HbA1c in the VIKING cohort (Supplementary Table 12). However, the association with HbA1c levels is not significant in CROATIA-Korcula, the cohort where we discovered the connection between *MSR1* and transferrin glycosylation, and it also does not replicate in ORCADES, suggesting that it might be a false positive association. We next checked whether any of the glycome-associated genes were significantly associated with health-related traits in UK Biobank. We used two repositories of aggregated rare-variants associations: Genebass^29^ and the AstraZeneca PheWAS portal^30^. Missense variants from the *MSR1* gene were significantly associated with insulin-like growth factor 1 levels (*IGF1*) in both Genebass (SKAT-O p-value = 4.6×10^−10^) and the PheWAS portal (p-value = 1.6×10^−24^, for the “ptv5pcnt” collapsing model).

## Discussion

Statistical power to detect associations with rare genetic variants can be increased by aggregating the association signals across multiple rare variants in a gene^31^, or by using genetically isolated populations where, due to genetic drift, some variants are increased in frequency compared to a general population^32^. Further, intermediate phenotypes, more proximal to the genes and consequently more strongly influenced by them, can be used as “proxies’ of complex diseases to boost power. Glycosylation, one of the most common post-translational modifications, is one such intermediate phenotype and has been implicated in many diseases^10,13,14^. Here, we utilised the power of genetic isolates, aggregation of multiple rare variants and intermediate phenotypes to study the effect of rare variants on glycosylation of two proteins and their effect on disease.

We performed multiple gene-based aggregation tests to assess associations with transferrin (N = 1907) and IgG (N = 4912) glycan traits in three isolated cohorts of European descent, testing rare (MAF<5%) pLoF and missense variants from whole exome sequencing. We found rare variants from 8 genes contributing to glycan levels of either IgG or transferrin. As previously observed in GWAS using imputed genotypes, transferrin and IgG glycans showed mostly protein-specific gene-based associations^18^, including genes encoding known glycosylation enzymes (transferrin - *TIRAP*, a gene in the proximity of *ST3GAL4*; IgG - *ST6GAL1* and *MGAT3*), transcription factors (transferrin - *FOXI1*), as well as other genes (transferrin - *MSR1*; IgG - *RFXAP*). On the other hand, rare variants in *FUT8* and *FUT6*, genes encoding fucosyltransferase enzymes adding core and antennary fucose structures to the synthesised glycan, were associated with glycosylation of both proteins. Previously we showed that, while glycosylation of both transferrin and IgG proteins is associated with genes encoding FUT6 and FUT8 fucosylation enzymes, these associations are driven by independent, protein-specific variants mapped to the regulatory region of the two genes^18^. Accordingly, here we identified rare variants in the exonic portions of *FUT8* and *FUT6*, acting independently or in concert with GWAS-identifiable variants.

We successfully replicated 4 gene-glycan associations (*FUT6, FUT8, TIRAP* and *MGAT3*); however, noting variants in certain genes were lower in frequency (*MSR1* and *FOXI1*) or completely absent (*ST6GAL1* and *RFXAP*) in replication cohorts, we were underpowered to replicate the glycan associations with the remaining four genes. Two of the 8 identified associations, the ones with variants from the *FUT8* and *MGAT3* genes, were driven by multiple rare variants simultaneously contributing to protein glycosylation. The association with variants from the *ST6GAL1* gene would have been discovered using single-point ExWAS (but not GWAS). Interestingly, for all three of these genes, we have also detected common variants independently affecting IgG and transferrin glycans. While four associations (*TIRAP, FUT6, MSR1* and *FOXI1*)could have been discovered using a GWAS of imputed genotype data, three of them (*FUT6, MSR1* and *FOXI1*) were with low frequency variants (0.02 < MAF < 0.05). The associations with the *RFXAP* gene could not have been discovered by either GWAS or ExWAS single-point analysis.

Except for *RFXAP* and *TIRAP*, all of 8 identified genes have already been associated with IgG and transferrin glycosylation in previous GWAS studies^17,18,20,22,23^. The novel gene *TIRAP* is located in close proximity to *ST3GAL4*, another glycosyltransferase-coding gene known to be associated with transferrin glycosylation. *TIRAP* has a function in the innate immune system, where it is involved in cytokine secretion and the inflammatory response^33,34^. The lead rare variant in the mask, rs8177399 (Supplementary Table 3), in addition to being an expression QTL (eQTL) for *TIRAP* and several other genes, is also a splicing QTL (sQTL) for *ST3GAL4* in whole blood (GTEx^35^, p-value = 1.9×10^−8^). The regulatory factor X-associated protein encoded by *RFXAP* gene, whose variants are associated with IgG glycans, is part of a multimeric complex, called the RFX DNA-binding complex, that binds to certain major histocompatibility (MHC) class II gene promoters and activates their transcription. MHC-II molecules are transmembrane proteins, found on the surface of professional antigen-presenting cells (including B cells)^36^, which have a central role in development and control of the immune response. While the mechanism of *TIRAP*’s influence on the glycome could be through controlling the splicing of the known glycosyltransferase enzyme ST3GAL4, the precise role of *RFXAP* in protein glycosylation still needs to be established.

Changes in the glycosylation patterns are often observed in a wide range of pathological states, such as cancer, inflammatory, autoimmune, neurodegenerative and cardiovascular diseases^37–40^. We thus assessed the potential involvement of glycome-associated genes in health, by performing, in the same three cohorts, gene-based association tests of 116 quantitative health-related traits, limited to genes whose rare variants we found associated with the protein glycomes. However, given the likely small effect-size of variants on complex diseases, we did not find any significant associations. On the other hand, using publicly available repositories of gene-based associations in the UK Biobank data, we found that rare missense variants from *MSR1* (associated with transferrin glycosylation) were also associated with blood levels of insulin-like growth factor 1 (IGF1). IGF1 is a hormone with significant structural and functional similarities to insulin: lower levels of IGF1 are associated with higher risk of Type 1 and 2 diabetes mellitus^41,42^. Recently, a rare deleterious missense variant in *IGF1* receptor (*IGF1R*) was found to be significantly associated with Type 2 diabetes in UK Biobank, further corroborating the link between IGF1 and diabetes^43^. In addition, genetic variants in *MSR1* have been previously associated with plasma levels of the galectin-3-binding protein^44^. Similarly to IGF1, galectin-3 has been identified as a marker and a pathogenic factor in type 2 diabetes, with the serum protein levels increased in type 2 diabetes patients^47–51^. An important part of iron delivery depends on recycling transferrin via clathrin-mediated endocythosis. Interestingly, binding of galectin-3 to transferrin can affect its intracellular trafficking^45,46^. Based on the glycosylation profile, galectin-3 was found bound only to a select, minor fraction (∼5%) of transferrin, while interestingly none or little was bound to IgG^46^. Overall, variants from the *MSR1* gene seem to have a pleiotropic effect on transferrin glycosylation, galectin-3 and IGF1. In turn, both galectin-3 and IGF1 are associated with type 2 diabetes. The potential role of glycosylation of transferrin in these processes still needs to be established.

In conclusion, we identified rare pLoF and missense variants associated with transferrin and IgG N-glycome, in both known and not previously reported genes (*TIRAP, RFXAP*). By utilising the power of genetic isolates and aggregated effects of rare variants, we discovered biologically relevant associations with a 300-fold up-drifted variant in the ORCADES cohort (in the sialyltransferase gene, *ST6GAL1*, affecting levels of sialylation of IgG) and associations independent of single-point GWAS and ExWAS analyses (in glycosyltransferase genes *FUT8* and *MGAT3*). Interestingly, many of glycan traits are influenced both by common and rare variants, revealing a complex genetic architecture of these intermediate phenotypes. While we did not find any robust links between glycome-associated genes and diseases in studied cohorts, we discover a potential link between transferrin glycosylation, galectin-3, IGF1 and diabetes. The exact mechanism behind these connections still needs to be confirmed and further explored. This study shows that, utilising the power of genetic isolates, gene-based aggregation tests and intermediate phenotypes such as glycosylation, rare variant associations are detectable even in relatively small sample sizes (low thousands). However, larger cohorts would be required to identify the contribution of rare variants to multifactorial, complex diseases.

## Methods

### Genotypic data

#### Exome sequencing

The “Goldilocks” exome sequence data for ORCADES, CROATIA-Korcula and VIKING cohorts was prepared at the Regeneron Genetics Center, following the protocol detailed in Van Hout *et al*.^2^ for the UK Biobank whole-exome sequencing project. In summary, the multiplexed samples were sequenced on the Illumina NovaSeq 6000 platform using S2 flow cells. The raw sequencing data was processed by automated analysis using the DNAnexus platform^52^, where files were converted to FASTQ format, and then aligned to GRCh38 genome reference using the BWA-mem^53^. Duplicated reads were identified and flagged by the Picard tool^54^. Genotypes for each individual sample were called using the WeCall variant caller^55^. During quality control, samples genetically identified as duplicates, showing disagreement between genetically determined and reported sex, high rates of heterozygosity or contamination, low sequence coverage (less than 80% of targeted bases achieving 20X coverage) or discordant with genotyping chip were excluded. The number of samples removed after quality control are listed in Supplementary Table 13 for each cohort. Finally, the “Goldilocks” dataset was generated by (i) filtering out genotypes with read depth lower than 7 reads, (ii) keeping variants having at least one heterozygous variant genotype with allele balance ratio greater than or equal to 15% (AB ≥ 0.15) or at least one homozygous variant genotype, and (iii) filtering out variants with more than 10% of missingness and HWE p<10^−6^. Overall, a total of 2,090 ORCADES (820 male and 1,270 female), 2,872 CROATIA-Korcula (1,065 male and 1,807 female) and 2,108 VIKING (843 male and 1,265 female) participants passed all exome sequence and genotype quality control thresholds. A pVCF file containing all samples passing quality control was then created using the GLnexus joint genotyping tool.^56^

#### Variant annotation

Exome sequencing variants were annotated as described in Van Hout, *et al*.^2^ In brief, each variant was labelled with the most severe consequence across all protein-coding transcripts, implemented using SnpEff^57^. Gene regions were defined according to Ensembl release 85. Variants annotated as stop gained, start lost, splice donor, splice acceptor, stop lost and frameshift were considered as predicted LOF variants. The deleteriousness of missense variants was assessed using the following algorithms and classifications (based on dbNSFP 3.2): (1) SIFT: “D” (Damaging), (2) Polyphen2_HDIV: “D” (Damaging) or “P” (Possibly damaging), (3) Polyphen2_HVAR: “D” (Damaging) or “P” (Possibly damaging), (4) LRT^58^: “D” (Deleterious) and (5) MutationTaster^59^: “A” (Disease causing automatic) or “D” (Disease causing). Missense variants were considered “likely deleterious” if predicted as deleterious by all five algorithms, “possibly deleterious” if predicted as deleterious by at least one of the algorithms and “likely benign” if not predicted as deleterious by any of the algorithms.

#### Generation of gene burden masks

For each gene, we grouped the variants in the gene in four categories (masks), based on severity of their functional consequence. Mask 1 included only predicted loss-of-function (pLoFs) variants, mask 2 consisted of pLoF variants and all missense variants, and masks 3 and 4 contained pLoF and predicted deleterious missense variants (“possibly deleterious” and “likely deleterious” for mask 3 and mask 4, respectively). We considered two separate variations of each mask based on the frequency of the minor allele of the variants that were screened in that group: MAF ≤ 5% and MAF ≤ 1%. Overall, up to 8 burden tests were performed for each gene (Supplementary Table 14). Consequently, the masks are not independent - certain masks will include the variants listed in a different mask and additional, less severe or more frequent variants.

#### Phenotypic data

##### Transferrin and IgG N-glycome quantification

Transferrin and total IgG N-glycome quantification for ORCADES, VIKING and CROATIA-Korcula samples was performed at Genos Glycobiology Laboratory, following the protocol described in Trbojević-Akmačić *et al*.^60^ for transferrin, in Pučić *et al*.^61^ for IgG in ORCADES cohort and batch 1 of CROATIA-Korcula cohort, in Trbojević-Akmačić *et al*.^62^ for IgG in VIKING cohort and batch 2 of CROATIA-Korcula cohort. In summary, proteins of interest were first isolated from blood plasma (IgG depleted blood plasma, in the case of transferrin) using affinity chromatography binding to anti-transferrin antibodies plates for transferrin and protein G plates for IgG. The protein isolation step was followed by release and labelling of N-glycans and clean-up procedure.. IgG N-glycans have been released from total IgG (all subclasses). N-glycans were then separated and quantified by hydrophilic interaction ultra-high-performance liquid chromatography (HILIC-UHPLC). As a result, transferrin and total IgG samples were separated into 35 (transferrin: TfGP1 − TfGP35) and 24 (IgG: GP1 − GP24) chromatographic peaks. It is worth noting that there is no correspondence structure-wise between transferrin TfGP and IgG GP traits labelled with the same number.

##### Normalisation and batch correction

Prior to genetic analysis, raw N-glycan UHPLC data was normalised and batch corrected to reduce the experimental variation in measurements. Total area normalisation was performed by dividing the area of each chromatographic peak (35 for transferrin, 24 for IgG) by the total area of the corresponding chromatogram. Due to the multiplicative nature of measurement error and right-skewness of glycan data, normalised glycan measurements were log10-transformed. Batch correction was then performed using the empirical Bayes approach implemented in the “ComBat” function of the “sva” R package^63^, modelling the technical source of variation (96-well plate number) as batch covariate. Batch corrected measurements were then exponentiated back to the original scale. Prior to further analysis, each glycan trait was rank transformed to normal distribution using the “rntransform” function from the “GenABEL” R package^64^.

##### Derived glycan traits

IgG derived traits analysed included those defined by Huffman *et al*.^65^, and were calculated using the glycanr R package. In addition, new derived traits were calculated for both transferrin and IgG, representing the overall presence of a certain sugar structure on the totality of transferrin/IgG N-glycan traits measured (e.g. percentage of fucosylation). These newly generated traits are expected to give a direct insight in the biological pathway involved in the addition of the sugar moiety to glycan structures. Exact formulas used for defining transferrin and IgG newly derived traits can be found in Supplementary Tables 15 and 16 respectively.

##### Health-related quantitative traits

To evaluate the potential effect of rare variants affecting glycome on health-related phenotypes, in the same cohorts we collected 148 health-related, quantitative traits (e.g. anthropological measurements, blood levels of proteins, metabolites and biomarkers). Excluding traits with fewer than 800 samples, a total of 116 traits were considered for analysis (75 traits for ORCADES, 79 for VIKING and 47 for CROATIA-Korcula cohort). Each health-related trait was rank transformed to normal distribution using the “rntransform” function from the “GenABEL” R package^64^, followed by applying the rare-variants association pipeline described below.

##### Gene-based aggregation analysis

We performed variant Set Mixed Model Association Tests (SMMAT)^66^ on rank-transformed glycan traits, fitting a GLMM adjusting for age, sex, sampling batch in the case of CROATIA-Korcula IgG glycan traits, and familial or cryptic relatedness by kinship matrix. The kinship matrix was estimated from the genotyped data using the ‘ibs’ function from GenABEL R package^64^. The SMMAT framework includes 4 variant aggregate tests: burden test, sequence kernel association test (SKAT), SKAT-O and SMMAT-E, a hybrid test combining the burden test and SKAT. The 4 variant aggregate tests were performed on 8 different pools of genetic variants, called “masks”, described above (Supplementary Table 14).

Discovery significance threshold was Bonferroni corrected for the approximate number of genes in the human genome, 20,000, and the number of independent glycan traits, 21 for IgG and 31 for transferrin (0.05/20000/31 = 8.06×10^−8^ for transferrin, 0.05/20000/21 = 1.19×10^−7^ for IgG). The number of independent glycan traits was estimated as the number of principal components that jointly explained 99% of the total variance of transferrin/IgG glycan traits in each cohort (Supplementary Tables 17 and 18). PCA was calculated on rank-transformed glycan traits, separately for each cohort, using the “prcomp” function from “factoextra” R package^67^. A gene association was considered significant if it passed the above-described Bonferroni corrected significance threshold in at least one of the 4 performed variant aggregate tests and if the cumulative allele count of the variants included in the gene was equal or higher than 10. Replication significance threshold was defined as P = 0.05 divided by the number of genes and independent glycans to be replicated. For IgG glycans, this threshold was P = 5.95×10^−4^ (P = 0.05/4 genes/21 glycans) and for transferrin glycans, this threshold was P = 3.23×10^−4^ (P = 0.05/5 genes/31 glycans).

A similar analysis plan was applied to the health-related phenotypes analysed. Variant Set Mixed Model Association Tests (SMMAT)^66^ was performed on rank-transformed traits, fitting a GLMM adjusting for age, sex, first 20 ancestral principal components (PCs), batch covariates when available (e.g. season, time of the day and batch/subcohort) and familial or cryptic relatedness.

##### IgG glycome gene-based aggregation meta-analysis

Gene-based aggregation analysis of IgG glycan traits for ORCADES and VIKING cohorts was repeated following the same approach as previously described, except for the restriction that masks included only variants present in both cohorts. Since IgG GP3 was not quantified in ORCADES cohort, this glycan was excluded from the meta-analysis, bringing the total number of IgG glycan traits considered to 93. We then used the “SMMAT.meta” function of “SMMAT” R package^66^ to meta-analyse, for each trait, the two studies. To identify significant results we filter results by the previously described Bonferroni-corrected significance threshold of 1.19×10^−7^ and by the cumulative allele count of variants included in the gene equal or higher than 10.

##### Genome-wide association analysis

Genome-wide association analyses (GWAS) between HRC-imputed genotypes and 51 transferrin N-glycan traits were performed in 948 samples from CROATIA-Korcula and 959 samples from VIKING. GWAS with 94 IgG N-glycan traits were performed in 1960 samples from ORCADES, 1866 samples from CROATIA-Korcula and 1086 samples from VIKING. The sample size of the same cohort differs between transferrin and IgG due to the different number of samples successfully measured for glycosylation of each protein. Transferrin N-glycan measurements were not available in ORCADES. Rank-transformed glycan traits were adjusted for age and sex, as fixed effects, and relatedness (estimated as the kinship matrix calculated from genotyped data) as random effect in a linear mixed model, calculated using the “polygenic” function from the “GenABEL” R package^64^. Since IgG N-glycan traits for the CROATIA-Korcula cohort were measured at two separate occasions, the two were considered as separate cohorts. Therefore, for CROATIA-Korcula, rank transformation was performed separately in each subcohort. Samples were then merged together for GWAS, but adding batch (subcohort number - 1 or 2) as fixed effect covariate. Residuals of covariate and relatedness correction were tested for association with Haplotype Reference Consortium (HRC) r1.1-imputed SNP dosages using the RegScan v. 0.5 software, applying an additive genetic model of association.

The genomic control inflation factor (λ_GC_) was calculated for each glycan and health-related trait. The mean genomic control inflation factor (λ_GC_) for IgG glycan traits was 1.002 (0.982-1.026) in ORCADES, 1 in CROATIA-Korcula (0.971-1.031) and 0.993 in VIKING cohort (0.972-1.017) cohort; for transferrin glycan traits λ_GC_ was 1.002 in CROATIA-Korcula (0.982-1.026) and 0.998 in VIKING (0.974-1.021) cohort. Overall, the confounding effects of the family structure were correctly accounted for in our analyses.

##### Identification of rare variant associations independent of GWAS and ExWAS signals

To ensure that the rare variant associations identified were independent of associations with variants discoverable by a GWAS or single-point exome-wide (ExWAS) analysis, we repeated the aggregate analysis while conditioning on the sentinel SNPs from the single-variant genome-wide or exome-wide analysis. First, we performed GWAS of glycan traits using the same individuals as in the analysis of the exome-sequencing data, but using as genotypes SNP dosages imputed from the HRC imputation panel, as described above. For each glycan trait we defined the sentinel SNPs as the variants having the lowest significant p-value (p < 5×10^−8^) in a 1Mb window, and MAF > 1%. Then we also performed the exome-wide association analysis (ExWAS), following exactly the same protocol, but with exome sequencing data used for genotypes. We then re-run variant aggregate analysis as previously described, but with adjusting the glycan traits for the genotype of the sentinel SNPs from the GWAS/ExWAS significant loci, in addition to the other covariates listed above. The statistical significance level was determined in the same way as outlined in the main analysis above.

##### Replication of glycome rare associations in different cohorts and associations with health-related traits

To investigate whether glycome rare-variants associations were cohort specific, each significant gene-glycan trait pair from the cohort-level discovery analysis was tested for associations in the remaining cohorts. The p-value threshold for replication was set to 3.23×10^−4^ for transferrin (0.05/31/5) and 5.95×10^−4^ for IgG (0.05/21/4) glycans, correcting for the number of independent glycan traits (i.e. 31 for transferrin and 21 for IgG) and the number of discovered glycome-gene pairs (i.e. 5 for transferrin and 4 for IgG in gene-based aggregation analysis).

To investigate whether the glycome associated rare-variants may also affect health-related phenotypes, we tested for association each glycome-associated gene and 116 health-related traits The significance threshold was set to 5.43×10^−5^, correcting for the number of health-related traits (116), and the number of discovered glycome-gene pairs and number of glycome-associated genes (8).

## Data Availability

There is neither Research Ethics Committee approval, nor consent from individual participants, to permit open release of the individual level research data underlying this study. The datasets generated and analysed during the current study are therefore not publicly available. Instead, the research data and/or DNA samples are available from on reasonable request, following approval by the QTL Data Access Committee and in line with the consent given by participants. Each approved project is subject to a data or materials transfer agreement (D/MTA) or commercial contract. The UK Biobank genotypic data used in this study were approved under application 19655, 48511 and 19655 are available to qualified researchers via the UK Biobank data access process. The expression data used for the analyses described in this manuscript were obtained from the GTEx Portal on 25/11/2022. Genebass (https://app.genebass.org/) and AstraZeneca PheWAS portal (https://azphewas.com/) were accessed on 06/12/2022.

## Code availability

We used publicly available software tools for all analyses. These software tools are listed in the main text and in the Methods.

## Acknowledgements

We thank Dr Nicola Pirastu for his help and advice regarding the statistical methods. The Orkney Complex Disease Study (ORCADES) was supported by the Chief Scientist Office of the Scottish Government (CZB/4/276 and CZB/4/710), the Royal Society, the MRC Human Genetics Unit, Arthritis Research UK and the European Union framework program 6 EUROSPAN project (contract number LSHG-CT-2006-018947). ORCADES DNA extractions and genotyping were performed at the Genetics Core of the Clinical Research Facility, University of Edinburgh. We would like to acknowledge the invaluable contributions of the research nurses in Orkney, the administrative team in Edinburgh, and the people of Orkney. The CROATIA-Korcula study was funded by grants from the MRC (United Kingdom), European Commission Framework 6 project EUROSPAN (contract number LSHG-CT-2006-018947), Croatian Science Foundation (grant 8875) and the Republic of Croatia Ministry of Science, Education and Sports (216-1080315-0302). Genotyping was performed in the Genetics Core of the Clinical Research Facility, University of Edinburgh. We would like to acknowledge all the staff of several institutions in Croatia that supported the CROATIA-Korcula fieldwork, including, but not limited to, the University of Split and Zagreb Medical Schools, Institute for Anthropological Research in Zagreb, and the Croatian Institute for Public Health in Split. The Viking Health Study-Shetland (VIKING) was supported by the MRC Human Genetics Unit quinquennial programme grant ‘QTL in Health and Disease’. DNA extractions and genotyping were performed at the Edinburgh Clinical Research Facility, University of Edinburgh. We would like to acknowledge the invaluable contributions of the research nurses in Shetland, the administrative team in Edinburgh and the people of Shetland. We acknowledge support from the European Union’s Horizon 2020 research and innovation programme IMforFUTURE (A.L. and A.F.-H.: H2020-MSCA-ITN/721815); the RCUK Innovation Fellowship from the National Productivity Investment Fund (L.K.: MR/R026408/1) and the MRC Human Genetics Unit programme grant, ‘QTL in Health and Disease’ (J.F.W. and C.H.: MC_UU_00007/10). Finally, this research has been conducted using data from the UK Biobank Resource (under application 26041, 48511 and 19655). For the purpose of open access, the author has applied a Creative Commons Attribution (CC BY) licence to any Author Accepted Manuscript version arising from this submission.

## Ethics

All studies were approved by local research ethics committees and all participants have given written informed consent. The ORCADES study was approved by the NHS Orkney Research Ethics Committee and the North of Scotland REC. The CROATIA-Korcula study was approved by the Ethics Committee of the Medical School, University of Split (approval ID: 2181-198-03-04/10-11-0008). The VIKING study was approved by the South East Scotland Research Ethics Committee, NHS Lothian (reference: 12/SS/0151).

## Author contributions

A.L.: Data analysis and interpretation, visualisation, writing—original draft preparation, writing—review and editing. P.R.H.J.T.: preparation of pipeline for gene-based aggregation test of rare variants, writing—review and editing. A.F.-H.: computation of new derived IgG glycan traits, data interpretation. I.T.-A.: Quantification of transferrin and IgG N-glycans, computation of derived transferrin glycan traits, writing—review and editing. F.V.: Glycan data quality control. T.P.: Quantification of transferrin and IgG N-glycans. G.T.: preparation, quality control and annotation of whole-exome sequencing data, writing—review and editing. A.R.S.: Funding. O.P.: Genomic and demographic data provider for CROATIA-Korcula cohort. C.H.: Genomic and demographic data provider for CROATIA-Korcula cohort. G.L.: Conceptualisation, glycan data provider, writing—review and editing. J.F.W.: Funding, conceptualisation, genomic and demographic data provider for ORCADES and VIKING cohort, supervision, data interpretation, writing—original draft preparation, writing—review and editing. L.K.: Conceptualisation, supervision, data interpretation, writing—original draft preparation, writing—review and editing.

## Competing interests

P.R.H.J.T. is an employee of BioAge Labs, Inc. G.T. and A.R.S. are full-time employees of Regeneron Genetics Center and receive salary, stock and stock options as compensation. G.L. is the founder and owner of Genos Ltd, a private research organisation that specialises in the high-throughput glycomic analysis and has several patents in this field. A.F.-H., I.T.-A., F.V., and T.P. are employees of Genos Ltd. L.K. is an employee of Humanity Inc., a company developing direct-to-consumer measures of biological ageing. All other authors declare no competing interests.

